# Racism and Racial Injustice During COVID-19: Impact on University Student Mental Health

**DOI:** 10.1101/2022.08.30.22279409

**Authors:** Laurence M. Boitet, Claire Estep, Lisa Schwiebert, Kalani Upshaw, Caro Wolfner, Amy Hutson Chatham, Sherilyn Garner, Angela Stowe, Robin Gaines Lanzi

## Abstract

The purpose of the study is to understand how undergraduate, graduate, and professional students were affected by the events of racial injustice and the COVID-19 pandemic. Data gathered from an online campus-wide survey administered during July and August 2020 indicated high levels of stress and rates of depression across all stages of training. A majority of these students also indicated that, while events around racism negatively impacted their mental health, such events did not affect students’ academic success as COVID-19 did. Although previous studies have demonstrated that student mental health has been negatively affected during COVID-19, this study shows that student mental health is also impacted by events driven by racism and racial injustice concurrent to the pandemic. In light of these findings, it is recommended that institutions adopt an intersectional approach toward addressing such contemporaneous stressors with initiatives that can adapt to multiple events simultaneously.

## Introduction

Federal and local responses were enacted immediately following the declaration of the COVID-19 public health emergency in January 2020. These included college and university campus shutdowns nationwide to minimize the spread of the virus, which incidently caused isolation and related hardships for students at all stages of training. Collectively, these actions have resulted in unprecedented psychological, social, and economic burdens with concomitant increases in stress, depression, and anxiety levels for these students (Ayittey et al., 2020; Lust et al., 2020; Son et al., 2020; Wenham et al., 2020). As reported recently by Stowe et al., students’ emotional and behavioral actions in response to COVID-19 can be described as an adaptive multi-phase process that aligns with Maslow’s Hierarchy of Needs (Stowe et al., 2021). The increase of student mental health concerns negatively impacted concentration and motivation, processes required for academic success, and increased engagement in negative coping strategies such as substance use (Gruber et al., 2021; Gueldner et al., 2020; Son et al., 2020; World Health Organization, 2020).

What is less recognized, however, is how the impact of racism and racial unrest during the COVID-19 public health emergency have further exacerbated student mental health, well-being, and coping strategies. While previous studies have demonstrated heightened mental health concerns (Priest et al., 2013) and a decreased use of mental health resources by students of color (Lipson et al., 2018), to our knowledge none have assessed the compond impact of racism and racial unrest on the stresses of a pandemic on their mental health and wellness.

To address this gap in knowledge, we assessed the impact of COVID-19 and racial unrest on students at a public university in the southeast. Our study surveyed undergraduate, graduate, and professional students to understand how these individuals were affected, both academically and personally, by these concurrent challenges. It is anticipated that results from this study will inform institutional strategies and initiatives to better support undergraduate, graduate, and professional students’ mental health during unprecedented events in the future.

## Methods

### Study Design and Sample Collection

Data were gathered via a campus-wide survey administered at a public, urban research-intensive university and academic health center with a large undergraduate college and nine professional schools in the southeastern US between July 7, 2020 and August 18, 2020. Recruitment materials (flyers with a QR code) were distributed through social media (Instagram, Facebook), emails, and campus-wide student organizations. A secured, online survey link and an informational sheet describing the study were provided to eligible participants. Eligible participants had to be enrolled as current UAB students (undergraduate, graduate, professional) and be willing and able to provide informed consent. This study was reviewed and approved by the Institutional Review Board.

### Measures

The survey contained a total of 62 quantitative and qualitative questions categorized into five separate sections: demographics, academics, COVID-19 impacts, racism and racial injustice impacts, and mental health. These sections included questions about demographics, stress, feelings, and impacts of COVID-19 and race-related events on academic success and mental health. A full accounting of measures’ results can be found in **Table 1**.

**Table 1.**
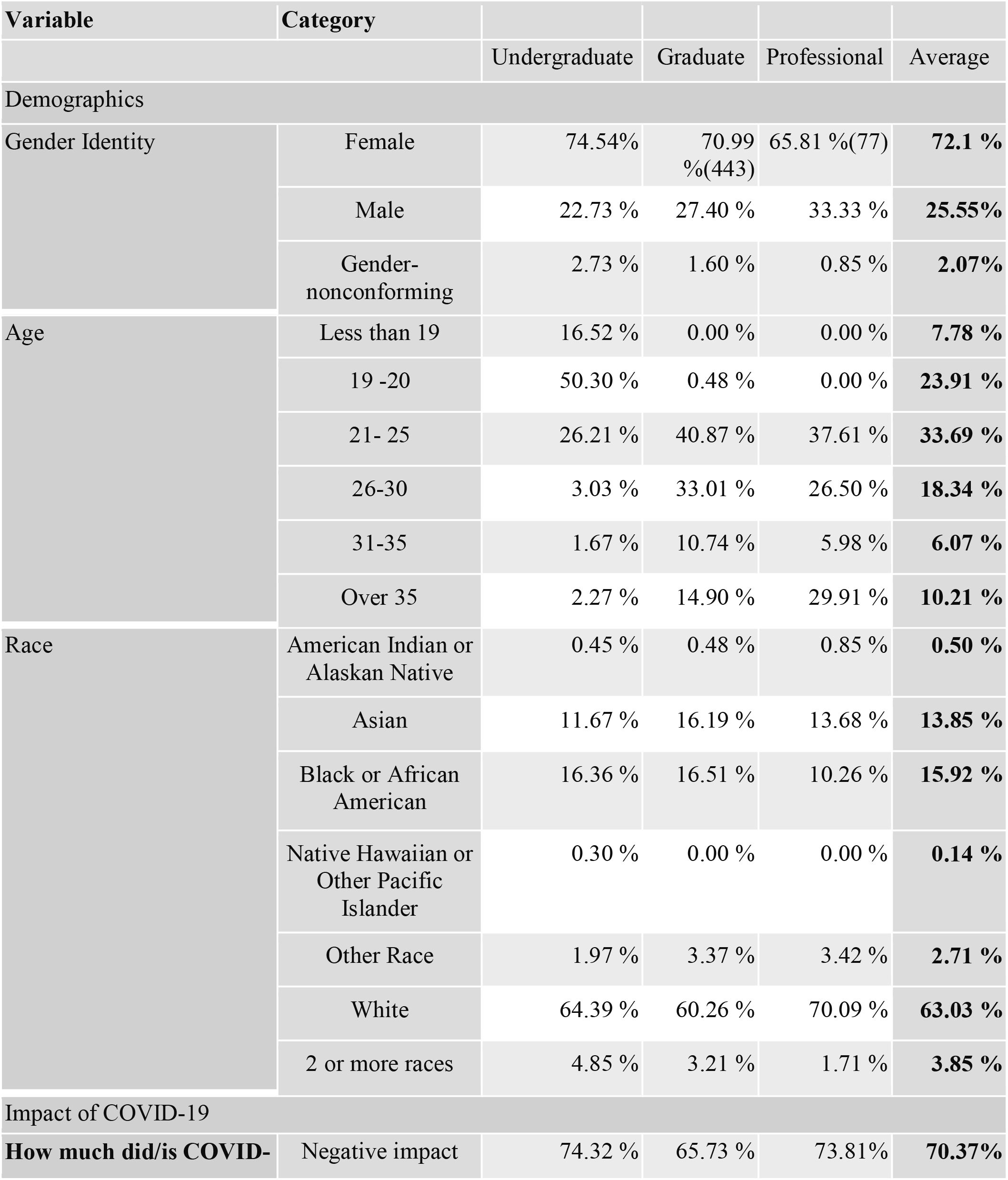
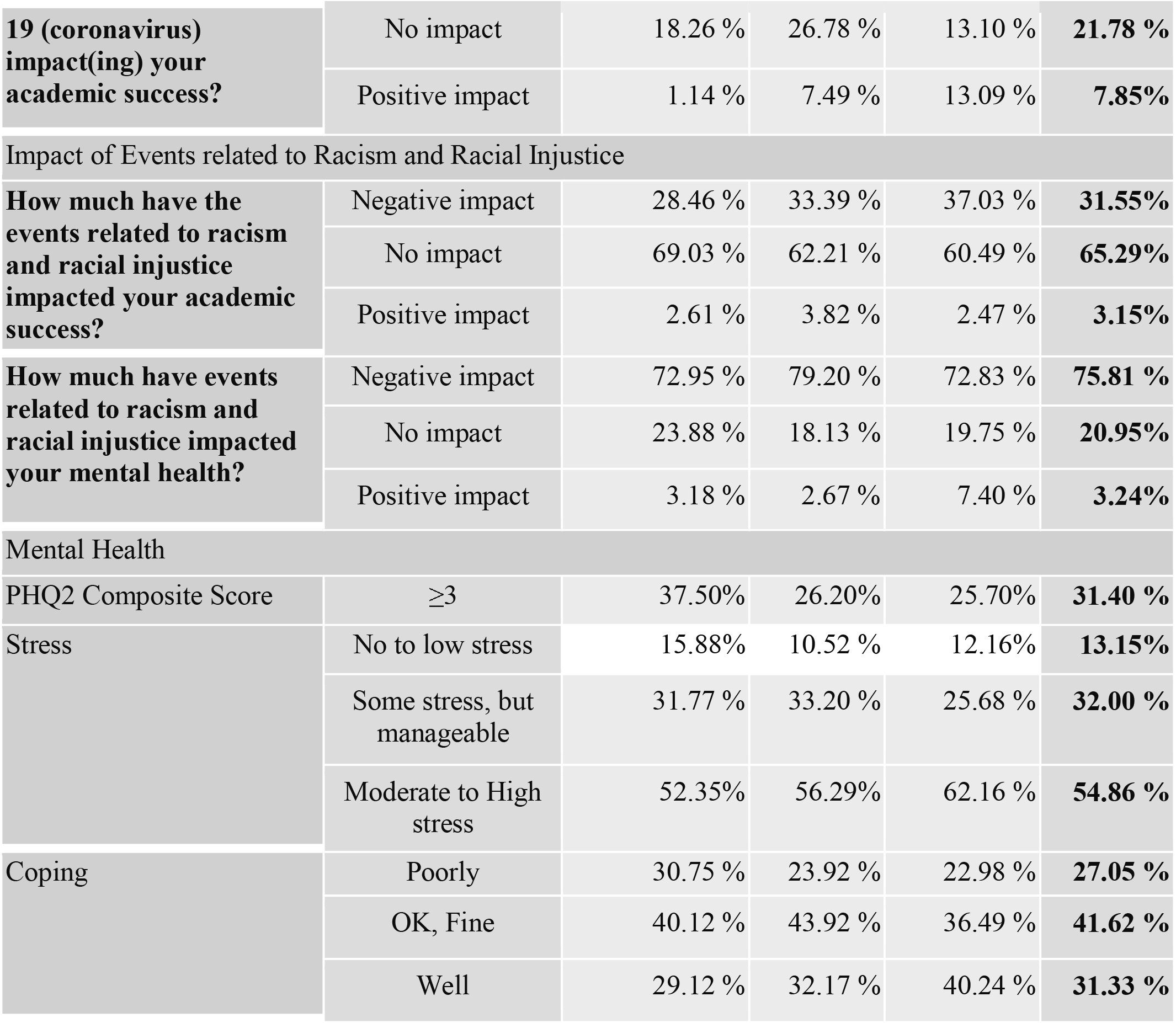
Survey Questionnaire Responses.

### Impacts of COVID-19 and Race-Related Events

To examine impacts of COVID-19, participants used a 5-point Likert scale to rate: “How much did/is COVID-19 (coronavirus) impact(ing) your academic success?”; and “How much did/is COVID-19 (coronavirus) impact(ing) your research?” (1, very negatively to 5, very positively). Additionally, to examine impacts of race-related events, participants used a 5-point Likert scale to rate: “How much have events related to racism and racial injustice impacted your mental health?”; “How much have the events related to racism and racial injustice impacted your academic success?”; “How much have events related to racism and racial injustice impacted your research?” (1, very negatively to 5, very positively).

### Patient Health Questionnaire-2

To assess depressive symptoms, participants completed the 2-tiem Patient Health Questionnaire-2 (PHQ-2) (α=0.9) measured on a 4-point Likert scale (0, not at all to 3, nearly every day) (Kroenke et al., 2001; Löwe et al., 2005; Richardson et al., 2010). Participants were asked how often they experienced little interest or pleasure in doing things and how often they experienced feeling down, depressed, or hopeless in the two weeks prior to taking the survey. A composite score was created from responses to these two items ranging from 0-6, where scores greater than or equal to 3 indicate significant depressive symptoms.

### Stress and Coping

To assess stress levels, participants were asked to “Rate your stress level” on a five point Likert scale (1, no stress to 5, high stress). In order to assess how students were coping, they were asked, on a five point Likert scale, to “Rate how you are coping currently” (1, extremely poorly to 5, extremely well). In addition, participants were asked “Select all that apply below to how you are currently feeling.*”* Participants were provided a list of 24 different feelings representing a range of emotions (e.g., frustration, anxiety, isolation, hope, happiness, content). They were also provided the opportunity to list additional feelings that were not included. Participants were also asked to “Check any of the following events that have impacted your mental health” from a list of 24 choices covering a range of commonly experienced events (e.g., killings of Black Americans, difficulty accessing internet, change or loss in income, lack of space to study and do schoolwork). They were also provided the option to list any other events that were not included.

## Results

### Participant Descriptives

Study participants were comprised of 1401 university students from across campus: undergraduate (n=660, 47.11%), graduate (n= 624, 44.54%), and professional (n=117, 8.35%). The majority of participants were female (undergraduate, 74.54%; graduate, 70.99%; professional, 65.81%), non-Hispanic (undergraduate, 93.79%; graduate, 95.19%; professional, 96.58%), and white (undergraduate, 64.39%; graduate, 60.26%; professional, 70.09%). A full description of participant demographics is listed in **Table 1**.

### Stress and Depression

We found that participants indicated high levels of stress and rates of depression at the time of survey. Across all groups, 54.9% of participants reported moderate to high stress levels (undergraduate, 52.35%; graduate, 56.3%; professional, 62.2%). Similarly, across all groups, 31.4% of participants screened positive for major depressive disorder (PHQ-2 score of 3 or greater) with the largest proportion among undergraduates (37.5%) followed by graduate students (25.2%), followed by professional students (25.7%) **Table 1**.

### Events Impacting Mental Health

We report that the majority of participants experienced a negative impact of COVID-19 on academic success (undergraduate, 74.32%; graduate 65.73%; professional, 73.81%; overall, 70.37%). Participants further indicated isolation due to social distancing (64%), killings of Black/African Americans (63%), and local/national protests related to racism (61%) as factors that impacted their mental health since the beginning of the COVID-19 pandemic. The majority of participants in reported that the then current events around racism and racial injustice negatively impacted their mental health (percentage negative impact: undergraduate, 72.95%; graduate, 79.20%; professional, 72.83%; overall, 75.81%). Interestingly, the participants did not report a negative impact of racism and racial injustice on their academic success as frequently, compared to that of the pandemic (percentage negative impact: undergraduate, 28.46 %, graduate, 33.39 %; professional, 37.03 %; overall, 31.55%) **Table 1**.

### Coping

To understand how students were dealing with stress, participants were asked to rate how they felt they were coping at the time of survey. While 27% of participants across all groups indicated they were coping poorly, 31% indicated they were coping well. Although 72.95% of the participants indicated they were coping fine to well, uncertainty (77%), anxiety (72%), and frustration (68%) were the top feelings reported across all groups at the time of survey completion **Table 1**.

## Discussion

Although previous studies have indicated that student mental health has been negatively impacted as a result of the COVID-19 pandemic (Copeland et al., 2021; Liu et al., 2020), the present study suggests that a decline in student well-being reaches beyond the pandemic itself. Here we describe that undergraduate, graduate and professional student mental health was negatively impacted by other co-occurring events, including the killings of Blacks and African Americans and the protests related to racial injustice. Participant depression screening results, stress levels, and reported top feelings of uncertainty, anxiety, and frustration together indicate that students across these stages of training are not faring well in the midst of the stresses of COVID-19 and surrounding events. This increased stress is of concern as this population is already at an increased risk for negative mental health and increased instances of suicidality (Active Minds, 2020).

The majority of study participants indicated that COVID-19 impacted their mental health and acedemic success. The majority of participants also indicated the impact of racism and racial injustice on their mental health. Interestingly, participants less frequently reported negative impacts of racism and racial injustce events on their academic success. These results are likely attributed in part to COVID-19-related restrictions and limitations on the learning environment, such as lack of in-person instruction, limited access to research and inconsistent child-care availability. We postulate that study participants may have not viewed their feelings of uncertainty, anxiety and frustration as affecting their ability to concentrate and stay motivated for academic success.

The launch of the current study just weeks following the killing of George Floyd and during the height of the subsequent racial protests while in the midst of the COVID-19 public health emergency was well timed to gain understanding of the impact of these co-occurring events on student mental health. Further, the launch of this study at a university in the southeastern US, which has a deep history of racial unrest and violence, afforded a unique opportunity for this analysis. The current study, however, was not without limitations. Data were gathered during the 2020 summer semester when there was not only a great amount of uncertainty about the pandemic and subsequent specifics of returning to classes in the fall, but fewer students were enrolled as compared with fall and spring semesters that year. This limited enrollment may explain the skewed student participation, which was comprised largely of white females. This result may also in part have an effect on the impact of racism and racial unrest on perceived academic success.

Future work regarding the impact of COVID-19 compounded with racism on student mental health is required. The engagement of a more diverse representation of students at all training levels, including but not limited to males, other underrepresented groups and non-US citizens is needed to gain a greater and more complete understanding of the combined impact of these co-occurring events on students. Because the combined effects of the COVID-19 pandemic and racial injustice in society has not been resolved, future work should also evaluate the long-term impact of these events on students’ mental health and well-being. Lastly, future studies should retrospectively assess how the federal and local actions in response to the COVID-19 pandemic and racial unrest contributed to student mental health to alleviate stressors and better support students’ academic success and well-being.

While much attention is paid to the impact of COVID-19 on student mental health, this study indicates that the interrelated nature of the COVID-19 pandemic and the racial justice movements of 2020 affected the mental health and academic performance of students at all surveyed stages of training. In light of these findings, it is recommended that institutions construct policies and procedures that are able to effectively address student stressors and account for the compound effect of both healthcare and societal crises.

## Disclosures

The authors have no relevant financial or non-financial competing interests to report.

## Data Availability

All data produced in the present study are available upon reasonable request to the authors

## Notes

### Competing Interest Statement

The authors have declared no competing interest.

### Funding Statement

This work was supported by internal funds from the School of Public Health Back of
the Envelope Award, Department of Cell, Developmental and Integrative Biology, Department
of Health Behavior, School of Public Health, and the Office of Service Learning and
Undergraduate Research, all at the University of Alabama at Birmingham

### Author Declarations

Ethics committee/IRB of the University of Alabama at Birminghamgave ethical approval for this work

